# Utility of the CHADS-P_2_A_2_RC score for estimating net adverse clinical events in chronic coronary syndrome patients without atrial fibrillation: insights from the CLIDAS-PCI

**DOI:** 10.1101/2025.03.21.25324437

**Authors:** Takenobu Shimada, Daiju Fukuda, Atsushi Shibata, Kenichiro Otsuka, Asahiro Ito, Hiroshi Okamura, Tetsuya Matoba, Takahide Kohro, Yusuke Oba, Tomoyuki Kabutoya, Yasushi Imai, Kazuomi Kario, Arihiro Kiyosue, Yoshiko Mizuno, Kotaro Nochioka, Masaharu Nakayama, Takamasa Iwai, Yoshihiro Miyamoto, Masanobu Ishii, Taishi Nakamura, Kenichi Tsujita, Hisahiko Sato, Naoyuki Akashi, Hideo Fujita, Ryozo Nagai, CLIDAS Research Group

**Author notes:** Corresponding author: Takenobu Shimada, MD, PhD, Department of Cardiovascular Medicine, Osaka Metropolitan University Graduate School of Medicine, 1-4-3 Asahimachi Abenoku, Osaka 545-8585, Japan.

## Abstract

**Background:** There are few data verifying the utility of the CHADS-P_2_A_2_RC score in comparison with the CHADS_2_ score for estimating net adverse clinical events (NACE) in real-world settings.

**Methods:** We performed analysis for a total of 3,985 chronic coronary syndrome (CCS) patients without atrial fibrillation (AF) who underwent percutaneous coronary intervention (PCI) between April 2013 and March 2019 for whom information was obtained from the CLIDAS (Clinical Deep Data Accumulation System)-PCI database. The primary endpoint was NACE defined as the composite of 3-point major adverse cardiovascular events (3P-MACE) (cardiovascular death, non-fatal myocardial infarction, and non-fatal stroke) and GUSTO moderate/severe bleeding events.

**Results:** Kaplan-Meier analysis showed that both the CHADS-P_2_A_2_RC and CHADS_2_ scores stratified the risks. The incidences of NACE were stratified well by the very-high-risk category, which was uniquely defined as a CHADS-P_2_A_2_RC score of ≥6 (hazard ratio: 2.38, 95% CI=1.91-2.97, p-value <0.001). The area under the curve (AUC) in estimating NACE within 3 years was higher when the CHADS-P_2_A_2_RC score was used than when the CHADS_2_ score was used (0.67 vs. 0.62, p=0.003). This was mainly due to the accuracy in estimating bleeding events (0.66 vs. 0.60, p=0.006).

**Conclusions:** The accuracy in estimating NACE after PCI for CCS patients without AF was higher when the CHADS-P_2_A_2_RC score was used than when the CHADS_2_ score was used, mainly due to the accuracy in predicting bleeding risk. Higher incidences of endpoints were well-stratified by a very-high-risk category defined as a CHADS-P_2_A_2_RC score of ≥6.

## Introduction

Administration of appropriate antithrombotic agents plays an important role in secondary prevention of coronary artery disease^1–4^. However, antithrombotic agents have the downside of increasing the risk of bleeding, and a simple risk model that can estimate net adverse clinical events (NACE) including both thrombotic and bleeding events is desirable.

The CHADS-P_2_A_2_RC score was originally developed as a prognostic prediction model for assessing the thrombotic events in patients without atrial fibrillation (AF)^5^. Recently, it was reported that the CHADS-P_2_A_2_RC score can be used for assessing NACE in patients with chronic coronary syndrome (CCS)^6,7^. The CHADS-P_2_A_2_RC score has been increasingly recognized as a criterion for initiation of treatment with a low-dose direct oral anticoagulant (DOAC) in patients with CCS^6^. However, there are still no data regarding its utility in real-world settings in comparison with other risk models.

The CHADS_2_ score was originally developed as a tool for evaluating the risk of stroke and it is widely used as a criterion for the initiation of anticoagulant therapy in patients with AF^8^. Recently, the CHADS_2_ score has also been shown to be useful for predicting prognosis in patients with CCS who do not have AF^9–11^.

In this study, to verify the utility of the CHADS-P_2_A_2_RC score for estimating NACE including thrombotic and bleeding events, we compared its accuracy with that of the CHADS_2_ score in CCS patients who underwent percutaneous coronary intervention (PCI) in real-world settings.

## Materials and methods

### Data source

The analysis in this study was conducted retrospectively using the Clinical Deep Data Accumulation System (CLIDAS)-PCI database that was formerly referred to as CLIDAS^12,13^, which was started as the Japan Ischemic heart disease Multimodal Prospective data Acquisition for preCision Treatment (J-IMPACT) project launched in 2015^14^. The database is a retrospective all-comer registry including real-world data for a total of 9,690 patients with acute coronary syndrome (ACS) or CCS who underwent PCI between April 2013 and March 2019 at seven tertiary facilities in Japan (six university hospitals and one national cardiovascular center). This database directly extracted real-world clinical data electronically from the hospital information system (HIS) of each participating facility in standardized data formats for clinical studies. Briefly, Standardized Structured Medical Information eXchange version 2 (SS-MIX2) standard storage was used to collect essential patient information, prescriptions, and laboratory data; SS-MIX2 extended storage was used to collect echocardiographic parameters, electrocardiograms, and cardiac catheterization reports^15^. Researchers and data managers at each facility collected the nonstandardized information on patients’ backgrounds and follow-up data. After data collection, each facility’s data were sent to the central server after anonymization^12–15^.

### Study design

The aim of this study was to determine the utility of CHADS-P_2_A_2_RC score in CCS patients without AF for estimating NACE in comparison with the CHADS_2_ score. The CHADS-P_2_A_2_RC score was originally developed in a cohort without AF^5^, and its utility was previously reported in a CCS cohort^6,7^. Thus, CCS patients without AF were also analyzed in this study. Patients lacking any data among the factors of the CHADS-P_2_A_2_RC or CHADS_2_ score were excluded. The study flow is shown in Figure 1.

**Figure 1.**
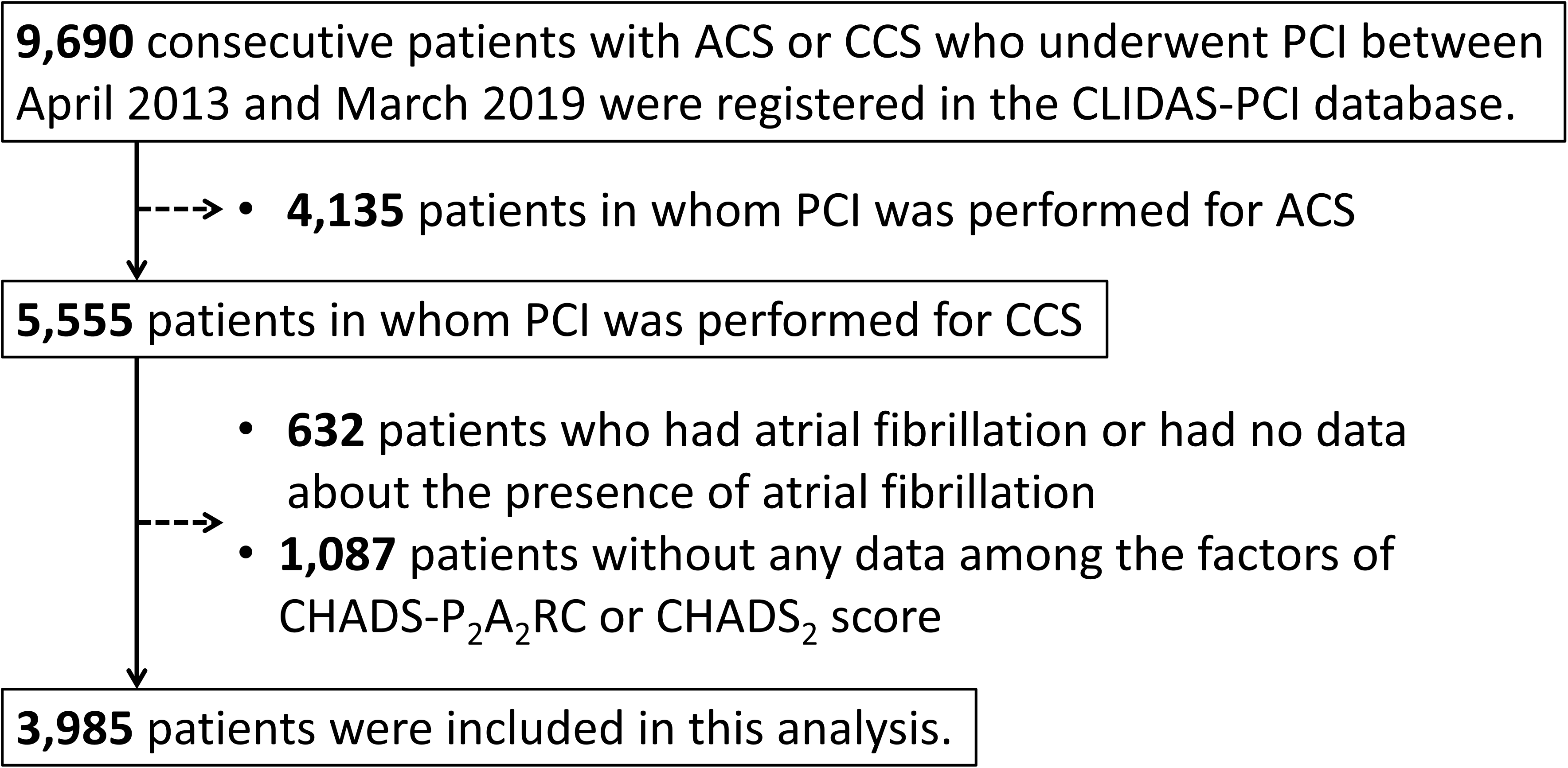
Study flowchart. Information for CCS patients without atrial fibrillation were obtained from the CLIDAS-PCI database. Abbreviations: ACS, acute coronary syndrome; CCS, chronic coronary syndrome; PCI, percutaneous coronary intervention; CLIDAS, Clinical Deep Data Accumulation System

In this study, we examined the baseline patient background characteristics and long-term adverse events according to the baseline calculated points using the CHADS-P_2_A_2_RC and CHADS_2_ scores. The accuracy of the CHADS-P_2_A_2_RC score and the accuracy of the CHADS_2_ score for estimating each endpoint were compared.

This study complied with the Declaration of Helsinki for investigation in human beings and was conducted with the approval of the Jichi Medical University Bioethics Committee for Medical Research (23-158). The requirement for written informed consent was waived owing to the retrospective nature of the study, and patients were allowed to opt out of the study alternatively.

### Risk stratification using the CHADS-P_2_A_2_RC and CHADS_2_ scores

The CHADS-P_2_A_2_RC score assigns one point each to congestive heart failure, hypertension, age of 65–74 years, diabetes mellitus, active smoking, chronic kidney disease (CKD), and multi-vessel coronary artery disease (CAD) and it assigns two points each to peripheral arterial disease (PAD) and age ≥75 years^5^. The CHADS_2_ score assigns one point each to congestive heart failure, hypertension, age ≥75 years, and diabetes mellitus and it assigns two points to stroke^8^.

Furthermore, we categorized the study patients according to the baseline CHADS-P_2_A_2_RC and CHADS_2_ scores. Classically, in the CHADS_2_ score, patients with scores of 0–1 were classified as patients with low risk, patients with scores of 2–3 were classified as patients with intermediate risk, and patients with scores of 4–6 were classified as patients with high risk^8^. In a recent study focusing on the CHADS-P_2_A_2_RC score, patients were classified in the same manner as that for the CHADS_2_ score: patients with scores of 0–1 were classified as patients with low risk, patients with scores of 2–3 were classified as patients with intermediate risk, and patients with 4 or higher were classified as patients with high risk^5^. However, the maximum CHADS-P_2_A_2_RC score is 11, we hypothesized that the high risk category for a CHADS-P_2_A_2_RC score of 4 or higher may include more patients than we want to identify who have a truly high risk for adverse events in real-world settings. Thus, we set a very-high-risk category uniquely based on the distribution of patients according to the points of the CHADS-P_2_A_2_RC scores.

### Endpoints and definitions

The primary endpoint was NACE defined as the composite of 3-point major adverse cardiovascular events (3P-MACE) and bleeding event. 3P-MACE were defined as the composite of cardiovascular death, non-fatal myocardial infarction, and non-fatal stroke. Bleeding event was defined as a moderate or severe bleeding event according to the Global Utilization of Streptokinase and t-PA for Occluded Coronary Arteries Trial (GUSTO) classification^16,17^. Briefly, moderate bleeding is bleeding that requires blood transfusion but does not result in hemodynamic compromise, and severe bleeding is either intracranial hemorrhage or bleeding that causes hemodynamic compromise and requires intervention. The secondary endpoints were 3P-MACE and bleeding event, which were components of the primary endpoint, and the 3-year cumulative incidence of each endpoint.

All baseline laboratory data were calculated as average values between 60 days before and 30 days after the index PCI. Estimated glomerular filtration rate (eGFR) was calculated by the Japanese equation using serum creatinine level, age, and sex as follows: eGFR (mL/min/1.73 m^2^) = 194 × Cr^−1.094^ × age^−0.287^ (× 0.739 for women)^18^. We defined CKD as eGFR <60 ml/min per 1.73 m^2 19^. PAD was defined as a history of non-cardiac (aortic or peripheral vascular) vascular disease. The number of diseased vessels was defined as the number of coronary arteries with severe stenosis (≥75%) in the major epicardial coronary segments of the right coronary artery, left anterior descending artery, and left circumflex artery, including their branch lesions, in which PCI was performed. The diseased left main trunk (LMT), defined as ≥75% stenosis, was counted separately. Multi-vessel CAD was defined as non-single vessel disease^13^.

### Statistical analysis

The baseline characteristics of the patients were summarized using medians [interquartile range (IQR)] for continuous variables and numbers (percentage) for categorical variables. For continuous data, the groups were compared using Wilcoxon’s rank-sum test. Categorical variables were compared using the chi-square test or Fisher’s exact test as appropriate.

The effect of each factor of the CHADS-P_2_A_2_RC and CHADS_2_ scores on each endpoint was analyzed using the Cox proportional hazard model and was expressed as the hazard ratio (HR) with 95% confidence interval (CI). The cumulative incidence of each endpoint was estimated by the Kaplan–Meier method and the cumulative incidences were compared by the log-rank test. The effect between each risk category was analyzed using the Cox proportional hazard model and was expressed as HR with the 95% CI. In the case of multiple comparisons, crude values were presented for interpretation using the Bonferroni method. To assess the model performance, we compared the areas under the receiver operating characteristic curves (AUC) by estimating the 3-year cumulative incidence of each endpoint.

All statistical analyses were conducted using R version 4.3.1 (R Foundation for Statistical Computing).

## Results

### Study population

A total of 3,985 CCS patients without AF who underwent PCI were included in this study. The baseline patient characteristics are shown in Table 1. There were high prevalences of comorbidities including hypertension in 83.6% of the patients, diabetes mellitus in 48.1% of the patients, CKD in 48.2% of the patients, PAD in 11.4% of the patients, and malignancy in 11.8% of the patients. Multi-vessel CAD present in 52.0% of the patients. Regarding lipid management, the median LDL-C level was 88.6 [IQR:71.8-107.6] and the statin administration rate was less than 80%, which did not meet the current guidelines’ recommendation. However, the administration rates of RAS inhibitors and β-blocker were more than 50%, which may be high for CCS patients. The median follow-up duration was 951 [IQR:328-1695] days.

**Table 1.**
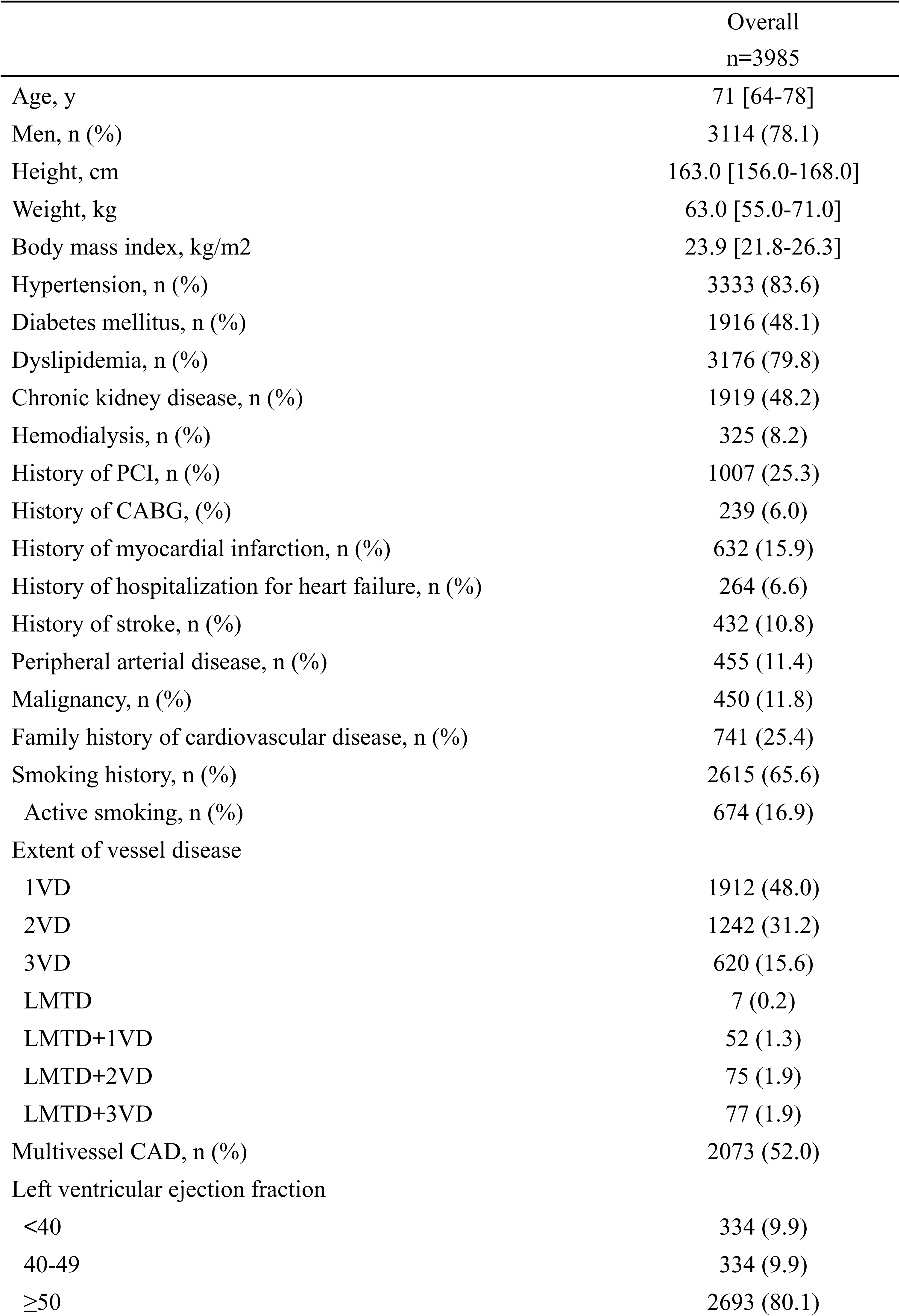
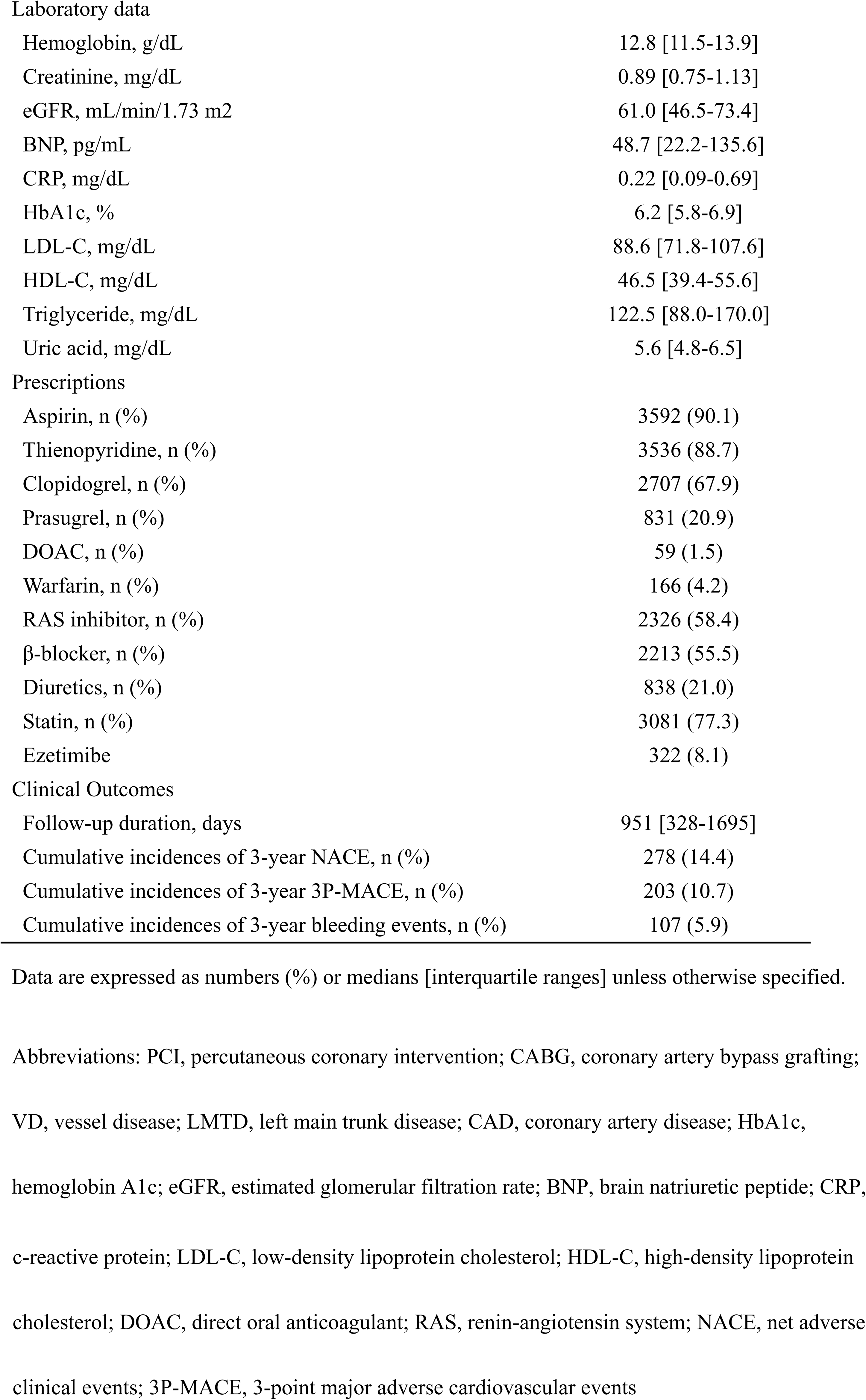
Baseline characteristics of the patient population.

Figure 2 shows the distributions of patients according to scores. The CHADS_2_ score seemed to appropriately categorize the population according to the original criteria, by which 1,363 patients (34.2%) with scores of 0–1 were categorized as patients with low risk, 2,255 patients (56.6%) with scores of 2–3 points were categorized as patients with intermediate risk, and 367 patients (9.2%) with scores of 4–6 points were categorized as patients with high risk. On the other hand, in the CHADS-P_2_A_2_RC score, if all scores of 4 or more were categorized as high risk, more than half of the patients would be categorized as patients with high risk. Thus, based on the distribution of patients with each score, we decided to categorize patients with a score of 6 points or more as patients with a very high risk. By this categorization, 277 patients (7.0%) with scores of 0–1 were categorized as patients with low risk, 1,333 patients (33.5%) with scores of 2–3 were categorized as patients with intermediate risk, 1,746 patients (43.8%) with scores of 4–5 were categorized as patients with high risk, and 629 patients (15.8%) with scores of 6-11 points were categorized as patients with very high risk.

**Figure 2.**
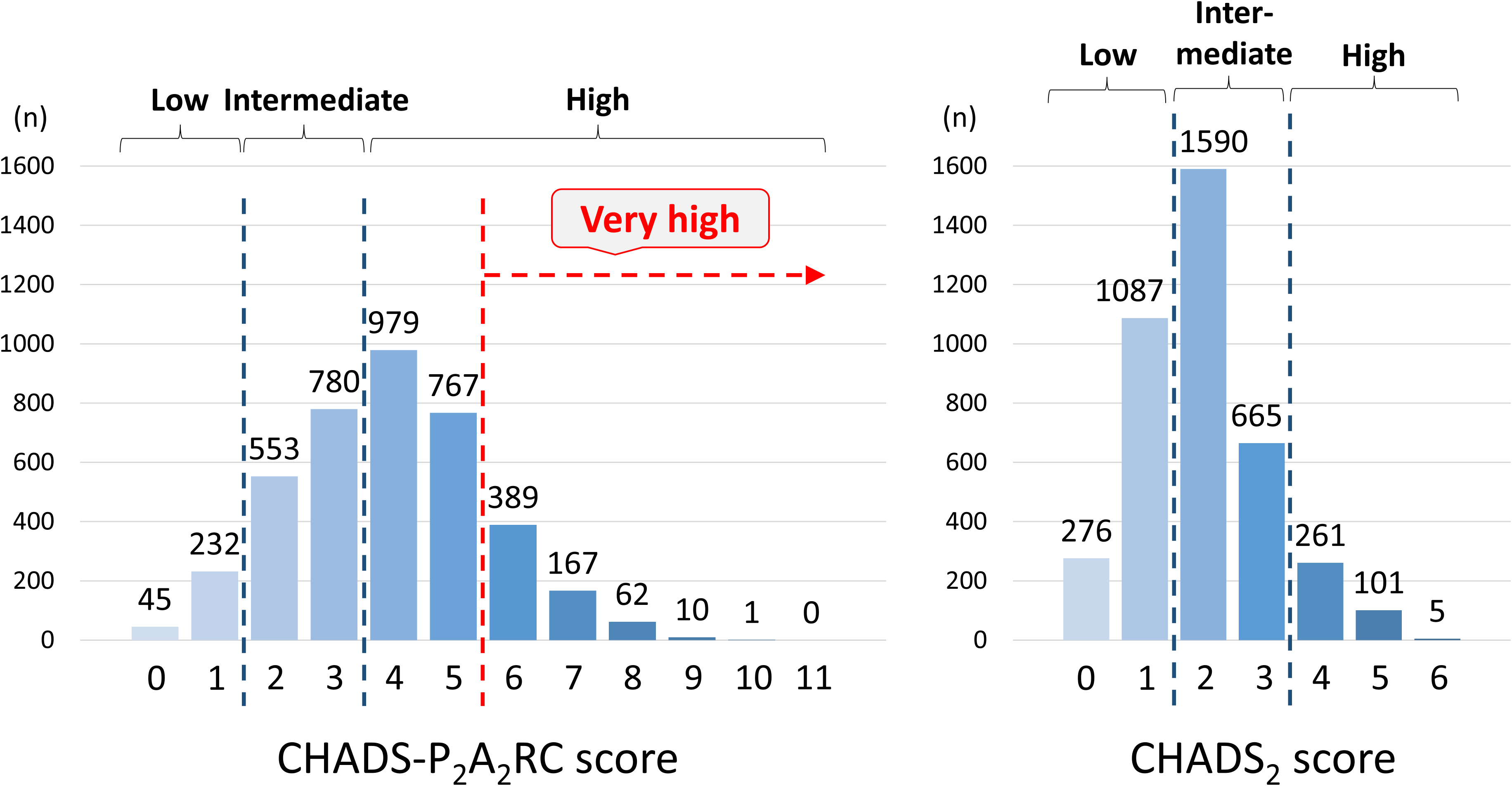
Distributions of patients according to the CHADS-P_2_A_2_RC and CHADS_2_ scores. The left side shows the numbers of patients categorized by the CHADS-P_2_A_2_RC scores, and the right side shows the numbers of patients categorized by the CHADS_2_ scores.

### Classification using CHADS-P_2_A_2_RC and CHADS_2_ scores

The baseline characteristics of patients in each category of CHADS-P_2_A_2_RC and CHADS_2_ scores are shown in Supplemental Table 1 and Supplemental Table 2, respectively. The backgrounds of patients in the categories differed significantly. The higher the risk categorization was, the higher was the proportion of poor background factors for all factors except for the LDL-C value and rate of statin administration. The percentage of patients with dyslipidemia was lowest in the low-risk category; however, LDL-C level and rate of statin administration were highest in the low-risk categories. The percentage of patients treated with clopidogrel was higher in the high-risk category, whereas the percentage of patients treated with prasugrel was higher in the low-risk category. These findings were the same for categories according to the CHADS-P_2_A_2_RC score and categories according to the CHADS_2_ score.

Table 2 shows the results of univariate Cox hazard regression analysis for NACE, 3P-MACE, and bleeding events according to the baseline factors of each CHADS-P_2_A_2_RC and CHADS_2_ score. Stroke and diabetes mellitus were not associated with bleeding events and the active smoking was not associated with each endpoint; however, all of the other factors were significantly associated with each endpoint.

**Table 2.** Cox hazard regression analyses for clinical outcomes according to baseline factors.

|  | NACE |  |  | 3P-MACE |  |  | Bleeding events |  |  |
| --- | --- | --- | --- | --- | --- | --- | --- | --- | --- |
|  | HR | 95% CI | p-value | HR | 95% CI | p-value | HR | 95% CI | p-value |
| Heart failure | 2.65 | 2.01-3.50 | <0.001 | 2.89 | 2.11-3.95 | <0.001 | 2.091 | 1.29-3.38 | 0.003 |
| Hypertension | 1.66 | 1.19-2.32 | 0.003 | 1.71 | 1.15-2.54 | 0.008 | 1.65 | 0.97-2.81 | 0.07 |
| Diabetes mellitus | 1.39 | 1.13-1.70 | 0.001 | 1.43 | 1.13-1.81 | 0.003 | 1.263 | 0.92-1.74 | 0.15 |
| Renal disease | 2.35 | 1.89-2.91 | <0.001 | 2.50 | 1.94-3.22 | <0.001 | 2.189 | 1.57-3.06 | <0.001 |
| Age ≥65 | 1.60 | 1.23-2.08 | <0.001 | 1.45 | 1.08-1.95 | 0.01 | 1.618 | 1.07-2.46 | 0.02 |
| Active smoking | 0.90 | 0.68-1.20 | 0.47 | 0.84 | 0.60-1.18 | 0.32 | 1.043 | 0.68-1.60 | 0.85 |
| Multivessel CAD | 1.53 | 1.24-1.89 | <0.001 | 1.57 | 1.23-2.01 | <0.001 | 1.364 | 0.98-1.90 | 0.07 |
| Age ≥75 | 1.75 | 1.43-2.14 | <0.001 | 1.83 | 1.44-2.31 | <0.001 | 1.673 | 1.21-2.31 | 0.002 |
| PAD | 1.80 | 1.39-2.33 | <0.001 | 1.54 | 1.13-2.11 | 0.007 | 1.939 | 1.30-2.89 | 0.001 |
| Stroke | 1.69 | 1.29-2.20 | <0.001 | 1.92 | 1.43-2.58 | <0.001 | 1.281 | 0.81-2.03 | 0.29 |

Results of univariate cox hazard regression for NACE, 3P-MACE, and bleeding events according to the baseline factors of CHADS-P<sub>2</sub>A<sub>2</sub>RC and CHADS<sub>2</sub> scores.
Abbreviations: NACE, net adverse clinical events; 3P-MACE, 3-point major adverse cardiovascular events; HR, hazard ratio; CI, confidence interval; CAD, coronary artery disease; PAD, peripheral arterial disease

### Impact of CHADS-P_2_A_2_RC and CHADS_2_ scores on each endpoint

Kaplan-Meier curves for each endpoint according to baseline categories of CHADS-P_2_A_2_RC and CHADS_2_ scores are shown in Figure 3. There were significant differences in all endpoints according to the categorization by both the CHADS-P_2_A_2_RC score and CHADS_2_ score. According to the features of these Kaplan-Meier curves, the incidences of endpoints were stratified well by the very-high-risk category according to the CHADS-P_2_A_2_RC score; however, the incidences of endpoints were not stratified well by the low-risk category according to the CHADS-P_2_A_2_RC score. On the other hand, the incidences of endpoints were stratified well by the low-risk category according to the CHADS_2_ score. In the very-high-risk category according to the CHADS-P_2_A_2_RC score, the rates of NACE were 7.4% (95% CI=5.5-9.9%) at 1 year, 19.0% (95% CI=15.6-23.1%) at 3 years and 30.8% (95% CI=25.6-36.8%) at 6 years after the index PCI. In the low-risk category according to the CHADS_2_ score, the rates of NACE were 2.0% (95% CI=1.4-3.0%) at 1 year, 4.6% (95% CI=3.4-6.2%) at 3 years and 9.9% (95% CI=7.5-13.0%) at 6 years after the index PCI. In the very-high-risk category according to the CHADS-P_2_A_2_RC score, in comparison with the other categories, there were significant differences in NACE (HR:2.38, 95% CI=1.91-2.97, p-value <0.001), 3P-MACE (HR:2.33, 95% CI=1.81-3.01, p-value <0.001), and bleeding events (HR:2.11, 95% CI=1.48-3.01, p-value <0.001). In the low-risk category according to the CHADS_2_ score, in comparison with the other categories, there were significant differences in NACE (HR:0.41, 95% CI=0.32-0.54, p-value <0.001), 3P-MACE (HR:0.36, 95% CI=0.27-0.50, p-value <0.001), and bleeding events (HR:0.52, 95% CI=0.35-0.77, p-value <0.001).

**Figure 3.**
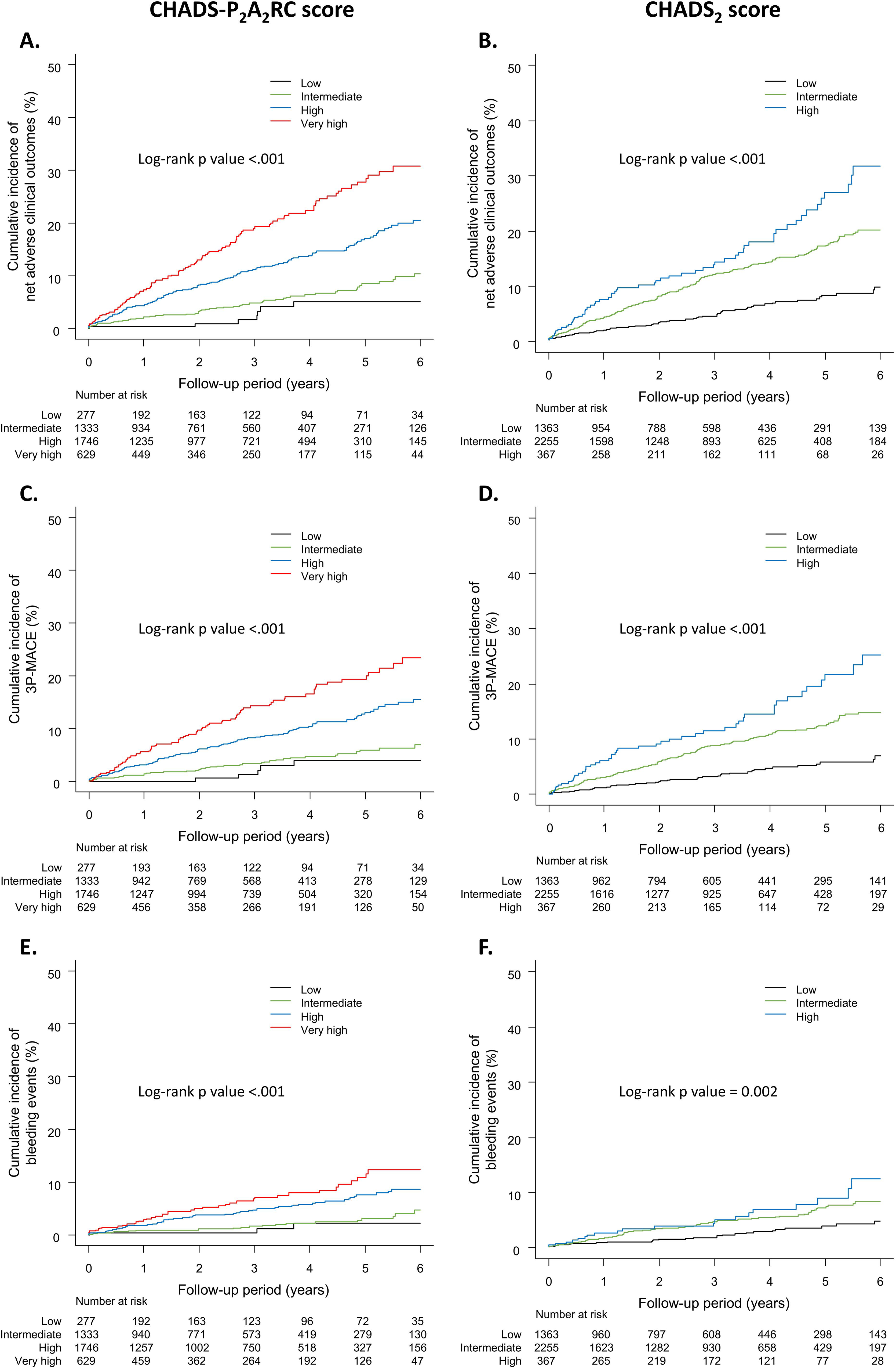
Kaplan-Meier curves for endpoints according to the baseline categorization by CHADS-P_2_A_2_RC and CHADS_2_ scores. A and B, Kaplan-Meier curves for NACE; C and D, Kaplan-Meier curves for 3P-MACE; E and F, Kaplan-Meier curves for bleeding events. The left side shows Kaplan-Meier curves according to the CHADS-P_2_A_2_RC score and the right side shows Kaplan-Meier curves according to the CHADS_2_ score. Abbreviations: NACE, net adverse clinical events; 3P-MACE, 3-point major adverse cardiovascular events

Hazard ratios analyzed in comparison with the low-risk category according to both the CHADS-P_2_A_2_RC score and CHADS_2_ score are shown in Supplemental Table 3. This analysis revealed that the low-risk category based on the CHADS-P_2_A_2_RC showed no significant differences from the intermediate-risk category in any endpoints.

### Accuracy of CHADS-P_2_A_2_RC and CHADS_2_ scores for estimating each endpoint

The results of ROC analyses for 3-year cumulative incidences of endpoints according to the CHADS-P_2_A_2_RC and CHADS_2_ scores are shown in Figure 4. The AUC in estimating NACE was higher when the CHADS-P_2_A_2_RC score was used than when the CHADS_2_ score was used; this was mainly attributed to the accuracy in estimating bleeding events with no significant difference in estimation of 3P-MACE.

**Figure 4.**
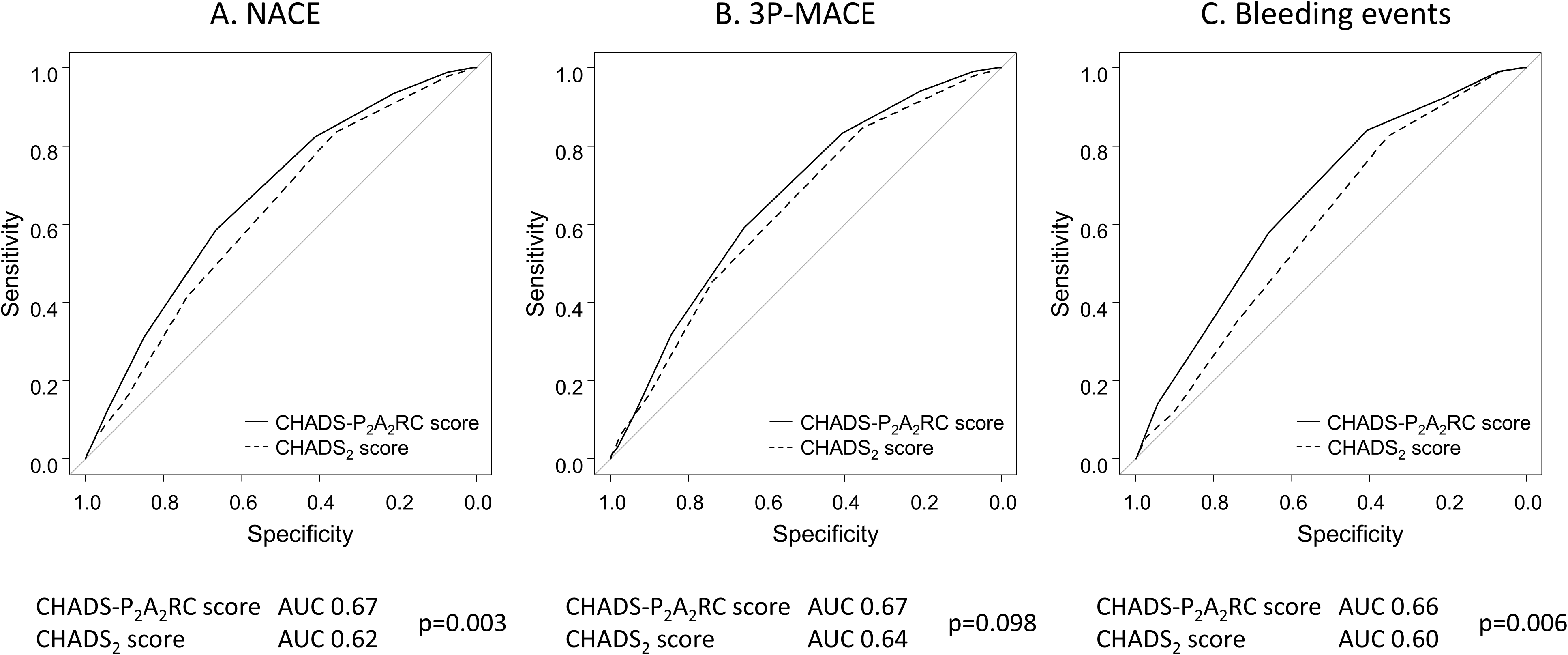
Receiver operating characteristic curve analysis of 3-year cumulative incidences of endpoints. A, NACE. B, 3P-MACE, and C: Bleeding events. Abbreviations: NACE, net adverse clinical events; 3P-MACE, 3-point major adverse cardiovascular events; AUC, area under curve.

## Discussion

The main findings of this study were as follows: (1) the risks for NACE including 3P-MACE and bleeding events were stratified well by both the CHADS-P_2_A_2_RC and CHADS_2_ scores, (2) higher incidences of endpoints were stratified well by a very-high-risk category defined as a CHADS-P_2_A_2_RC score of ≥6, whereas lower incidences of endpoints were stratified well by the low-risk category of the CHADS_2_ score, and (3) CHADS-P_2_A_2_RC score was superior to CHADS_2_ score for estimating bleeding events and was therefore superior for estimating NACE. In this study, we evaluated the utility of the CHADS-P_2_A_2_RC score in comparison with the CHADS_2_ score in real-world CCS patients without AF over a long-term period with a median follow-up period of 951 [IQR:328-1695] days.

In terms of the utility of the CHADS-P_2_A_2_RC score, the results of this study were consistent with the results of previous studies showing that the CHADS-P_2_A_2_RC score can risk-stratify NACE^6,7^. In this study, we set a new very-high-risk category defined as a CHADS-P_2_A_2_RC score of ≥6, and patients in that category had high incidences of NACE. Supplemental Figure 1 shows Kaplan-Meier curves when the very-high-risk category was defined as a CHADS-P_2_A_2_RC score of ≥7. According to the features of these Kaplan-Meier curves, the risk stratification of 3P-MACE was better when the very-high-risk category was defined as a CHADS-P_2_A_2_RC score of ≥6 than when it was defined as a CHADS-P_2_A_2_RC score of ≥7. Furthermore, 629 patients (15.8%) were categorized as patients with a very high risk when the cut-off of the CHADS-P_2_A_2_RC score was set as ≥6, and only 240 patients (6.0%) were categorized as patients with a very high risk when the cut-off of the CHADS-P_2_A_2_RC score was set as ≥7. If the prognosis can be improved by performing some sort of intervention without increasing the risk of bleeding, it is better to include more patients. Thus, we believe that the very-high-risk category defined as a CHADS-P_2_A_2_RC score of ≥6 is more appropriate than that defined as a CHADS-P_2_A_2_RC score of ≥7.

There are some well-known models for estimating both thrombotic and bleeding risks such as DAPT score^20^, PARIS score^21^, and CREDO-Kyoto risk score^22^, and there are some well-known models for estimating bleeding risk such as ARC-HBR criteria^23^ and PRECISE-DAPT score^24^. Among these risk models, DAPT score is a model that can estimate both thrombotic and bleeding risks from a single calculation. However, the factors of the DAPT score include uncommon factors such as the use of paclitaxel-eluting stents and vein graft intervention, and it may not be a practical model^20^. The CHADS-P_2_A_2_RC score does not require any special factors and is easy to use in daily clinical settings. Among the factors of the CHADS-P_2_A_2_RC and CHADS_2_ scores, CKD, active smoking, multi-vessel CAD, and PAD are factors that are included only in the CHADS-P_2_A_2_RC score, and stroke is a factor that is included only in the CHADS_2_ score. CKD, active smoking, and PAD are included as factors in some of the well-known risk scores for estimating thrombotic or bleeding risk, and coronary complexity is also known to be associated with the cardiovascular outcomes^25–27^. On the other hand, stroke is only included as a minor criterion in the ARC-HBR criteria^23^. The difference in accuracy between the CHADS-P_2_A_2_RC score and the CHADS_2_ score may have been affected by how many of these factors were included.

In this study, the percentage of patients with dyslipidemia was lowest in the low-risk category, whereas the LDL-C level and rate of statin administration were highest in the low-risk category (Supplemental Table 1 and Supplemental Table 2). The reason of this phenomenon is unclear; however, it might be due to the nature of real-world lipid management in which young patients with high LDL-C levels prefer to receive strict lipid management and older patients with lower LDL-C levels tend to receive mild lipid management^28^. Currently, statin administration is recommended regardless of the presence of dyslipidemia, and the concept of “lower the better” is widely known^29^, so the results of this part may differ in current clinical practice. This might be due to the enrollment period and the nature of the non-randomized real-world observational cohort.

In this study, high-risk patients tended to prefer to receive clopidogrel and low-risk patients tended to prefer prasugrel (Supplemental Table 1 and Supplemental Table 2). It has been reported that administration of prasugrel reduced cardiovascular events but also increased bleeding events compared to administration of clopidogrel^30^. East Asian people have a particularly high risk for bleeding^31^, and clopidogrel might have been preferred for safety concerns in patients who were considered to have a high risk for bleeding. On the other hand, administration of low-dose DOAC has been reported to reduce cardiovascular events while not significantly increasing major bleeding events^1^. Low-dose DOAC is not currently available in Japan, and future research focusing on the effectiveness and safety of low-dose DOAC in patients with a CHADS-P_2_A_2_RC score of ≥6 is needed.

## Limitations

There are several limitations in our study. First, our study was a retrospective study using data from a non-randomized observational registry. Since the calculations of CHADS-P_2_A_2_RC and CHADS_2_ scores were based on baseline information and the treatment provided to patients varied depending on the background of each patient, neither the baseline information nor the treatment provided could be adjusted, and all the results of this study were therefore just presented as crude data. Second, low-dose DOAC is not currently available in Japan, and the results of interventional analysis were therefore not included in this study. Only the utility and accuracy of the CHADS-P_2_A_2_RC and CHADS_2_ scores were assessed in this study. The utility of CHADS-P_2_A_2_RC score as a criterion for initiation of treatment with low-dose DOAC in real-world settings has not been verified, especially in East Asian patients, and future investigation is required. Third, the rate of statin administration was insufficient. This might be due to changes in recommendations for lipid management, and this may differ from current clinical practice. Fourth, the very-high-risk category was uniquely determined on the basis of the distribution of patients in this cohort, and the cutoff may be different in other patient cohorts.

## Conclusion

Long-term NACE in CCS patients without AF were stratified well by both the CHADS-P_2_A_2_RC and CHADS_2_ scores. The accuracy of risk stratification was higher for the CHADS-P_2_A_2_RC score than for the CHADS_2_ score due to the accuracy of the CHADS-P_2_A_2_RC score for predicting bleeding risk. Accurate identification of very-high-risk patients was achieved when the cut-off of the CHADS-P_2_A_2_RC score was set as ≥6.

## Data Availability

The data underlying this article will be shared on reasonable request to the corresponding author.

## Acknowledgements

This work was supported by the Cross-ministerial Strategic Innovation Promotion Program (SIP) on “Integrated Health Care System” Grant Number JPJ012425. In addition, the authors thank the Kowa Company for funding the development of CLIDAS-PCI. Finally, the authors appreciate the contributions of all CLIDAS research group members and the authors are grateful to Yuri Matoba (Precision Inc., Tokyo, Japan) for helping them integrate the data.

## References

1. Eikelboom JW, Connolly SJ, Bosch J, Dagenais GR, Hart RG, Shestakovska O, Diaz R, Alings M, Lonn EM, Anand SS, et al. Rivaroxaban with or without Aspirin in Stable Cardiovascular Disease. New England Journal of Medicine. 2017;377:1319–1330.

2. de Vries TI, Eikelboom JW, Bosch J, Westerink J, Dorresteijn JAN, Alings M, Dyal L, Berkowitz SD, van der Graaf Y, Fox KAA, et al. Estimating individual lifetime benefit and bleeding risk of adding rivaroxaban to aspirin for patients with stable cardiovascular disease: results from the COMPASS trial. Eur Heart J. 2019;40:3771–3778a.

3. Watanabe H, Morimoto T, Natsuaki M, Yamamoto K, Obayashi Y, Nishikawa R, Ando K, Ono K, Kadota K, Suwa S, et al. Clopidogrel vs Aspirin Monotherapy Beyond 1 Year After Percutaneous Coronary Intervention. J Am Coll Cardiol. 2024;83:17–31.

4. Natsuaki M, Watanabe H, Morimoto T, Yamamoto K, Obayashi Y, Nishikawa R, Ando K, Domei T, Suwa S, Ogita M, et al. An Aspirin-Free Versus Dual Antiplatelet Strategy for Coronary Stenting: STOPDAPT-3 Randomized Trial. Circulation. 2024;149:585–600.

5. Steensig K, Olesen KKW, Madsen M, Thim T, Jensen LO, Würtz M, Kristensen SD, Bøtker HE, Lip GYH, Eikelboom JW, et al. A Novel Model for Prediction of Thromboembolic and Cardiovascular Events in Patients Without Atrial Fibrillation. Am J Cardiol. 2020;131:40–48.

6. Würtz M, Olesen KKW, Mortensen MB, Eikelboom JW, Mohammad MA, Erlinge D, Kristensen SD, Maeng M. Dual antithrombotic treatment in chronic coronary syndrome: European Society of Cardiology criteria vs. CHADS-P2A2RC score. Eur Heart J. 2022;43:996–1004.

7. Würtz M, Olesen KKW, Bhatt DL, Yusuf S, Muehlhofer E, Eikelboom JW, Maeng M. Net clinical benefit of extended dual pathway inhibition according to baseline risk in patients with chronic coronary syndrome: a COMPASS substudy. Eur Heart J Cardiovasc Pharmacother. 2024;10:201–209.

8. Gage BF, Waterman AD, Shannon W, Boechler M, Rich MW, Radford MJ. Validation of Clinical Classification Schemes for Predicting Stroke. JAMA. 2001;285:2864.

9. Tabata N, Yamamoto E, Hokimoto S, Yamashita T, Sueta D, Takashio S, Arima Y, Izumiya Y, Kojima S, Kaikita K, et al. Prognostic Value of the CHADS _2_ Score for Adverse Cardiovascular Events in Coronary Artery Disease Patients Without Atrial Fibrillation—A Multi-Center Observational Cohort Study. J Am Heart Assoc. 2017;6. doi:10.1161/JAHA.117.006355.

10. Zhou X, Cao K, Kou S, QU S, Li H, Yu Y, Wang C, Liu Y, Li P, Li D. Usefulness of CHADS2 score for prognostic stratification of patients with coronary artery disease: A systematic review and meta-analysis of cohort studies. Int J Cardiol. 2017;228:906–911.

11. Sen J, Tonkin A, Varigos J, Fonguh S, Berkowitz SD, Yusuf S, Verhamme P, Vanassche T, Anand SS, Fox KAA, et al. Risk stratification of cardiovascular complications using CHA2DS2-VASc and CHADS2 scores in chronic atherosclerotic cardiovascular disease. Int J Cardiol. 2021;337:9–15.

12. Oba Y, Kabutoya T, Kohro T, Imai Y, Kario K, Sato H, Nochioka K, Nakayama M, Fujita H, Mizuno Y, et al. Relationships Among Heart Rate, β-Blocker Dosage, and Prognosis in Patients With Coronary Artery Disease in a Real-World Database Using a Multimodal Data Acquisition System. Circulation Journal. 2023;87:CJ-22-0314.

13. Akashi N, Matoba T, Kohro T, Oba Y, Kabutoya T, Imai Y, Kario K, Kiyosue A, Mizuno Y, Nochioka K, et al. Sex Differences in Long-Term Outcomes in Patients With Chronic Coronary Syndrome After Percutaneous Coronary Intervention ― Insights From a Japanese Real-World Database Using a Storage System ―. Circulation Journal. 2023;87:CJ-22-0653.

14. Matoba T, Kohro T, Fujita H, Nakayama M, Kiyosue A, Miyamoto Y, Nishimura K, Hashimoto H, Antoku Y, Nakashima N, et al. Architecture of the Japan Ischemic Heart Disease Multimodal Prospective Data Acquisition for Precision Treatment (J-IMPACT) System. Int Heart J. 2019;60:264–270.

15. Nakayama M, Takehana K, Kohro T, Matoba T, Tsutsui H, Nagai R. Standard Export Data Format for Extension Storage of Standardized Structured Medical Information Exchange. Circ Rep. 2020;2:587–616.

16. GUSTO investigators. An International Randomized Trial Comparing Four Thrombolytic Strategies for Acute Myocardial Infarction. New England Journal of Medicine. 1993;329:673–682.

17. Rao S V., O’Grady K, Pieper KS, Granger CB, Newby LK, Mahaffey KW, Moliterno DJ, Lincoff AM, Armstrong PW, Van de Werf F, et al. A Comparison of the Clinical Impact of Bleeding Measured by Two Different Classifications Among Patients With Acute Coronary Syndromes. J Am Coll Cardiol. 2006;47:809–816.

18. Matsuo S, Imai E, Horio M, Yasuda Y, Tomita K, Nitta K, Yamagata K, Tomino Y, Yokoyama H, Hishida A. Revised Equations for Estimated GFR From Serum Creatinine in Japan. American Journal of Kidney Diseases. 2009;53:982–992.

19. Stevens PE. Evaluation and Management of Chronic Kidney Disease: Synopsis of the Kidney Disease: Improving Global Outcomes 2012 Clinical Practice Guideline. Ann Intern Med. 2013;158:825.

20. Yeh RW, Secemsky EA, Kereiakes DJ, Normand S-LT, Gershlick AH, Cohen DJ, Spertus JA, Steg PG, Cutlip DE, Rinaldi MJ, et al. Development and Validation of a Prediction Rule for Benefit and Harm of Dual Antiplatelet Therapy Beyond 1 Year After Percutaneous Coronary Intervention. JAMA. 2016;315:1735.

21. Baber U, Mehran R, Giustino G, Cohen DJ, Henry TD, Sartori S, Ariti C, Litherland C, Dangas G, Gibson CM, et al. Coronary Thrombosis and Major Bleeding After PCI With Drug-Eluting Stents. J Am Coll Cardiol. 2016;67:2224–2234.

22. Natsuaki M, Morimoto T, Yamaji K, Watanabe H, Yoshikawa Y, Shiomi H, Nakagawa Y, Furukawa Y, Kadota K, Ando K, et al. Prediction of Thrombotic and Bleeding Events After Percutaneous Coronary Intervention: CREDO-Kyoto Thrombotic and Bleeding Risk Scores. J Am Heart Assoc. 2018;7. doi:10.1161/JAHA.118.008708.

23. Urban P, Mehran R, Colleran R, Angiolillo DJ, Byrne RA, Capodanno D, Cuisset T, Cutlip D, Eerdmans P, Eikelboom J, et al. Defining high bleeding risk in patients undergoing percutaneous coronary intervention: a consensus document from the Academic Research Consortium for High Bleeding Risk. Eur Heart J. 2019;40:2632–2653.

24. Costa F, van Klaveren D, James S, Heg D, Räber L, Feres F, Pilgrim T, Hong M-K, Kim H-S, Colombo A, et al. Derivation and validation of the predicting bleeding complications in patients undergoing stent implantation and subsequent dual antiplatelet therapy (PRECISE-DAPT) score: a pooled analysis of individual-patient datasets from clinical trials. The Lancet. 2017;389:1025–1034.

25. Sorajja P, Gersh BJ, Cox DA, McLaughlin MG, Zimetbaum P, Costantini C, Stuckey T, Tcheng JE, Mehran R, Lansky AJ, et al. Impact of multivessel disease on reperfusion success and clinical outcomes in patients undergoing primary percutaneous coronary intervention for acute myocardial infarction. Eur Heart J. 2007;28:1709–1716.

26. Head SJ, Davierwala PM, Serruys PW, Redwood SR, Colombo A, Mack MJ, Morice M-C, Holmes DR, Feldman TE, Stahle E, et al. Coronary artery bypass grafting vs. percutaneous coronary intervention for patients with three-vessel disease: final five-year follow-up of the SYNTAX trial. Eur Heart J. 2014;35:2821–2830.

27. Thuijs DJFM, Kappetein AP, Serruys PW, Mohr F-W, Morice M-C, Mack MJ, Holmes DR, Curzen N, Davierwala P, Noack T, et al. Percutaneous coronary intervention versus coronary artery bypass grafting in patients with three-vessel or left main coronary artery disease: 10-year follow-up of the multicentre randomised controlled SYNTAX trial. The Lancet. 2019;394:1325–1334.

28. Shimada T, Osakada K, Okabe K, Shima Y, Eguchi H, Habara S, Tada T, Taguchi Y, Ikuta A, Takamatsu M, et al. Impact of high-dose statin on cardiovascular outcomes in real-world patients with ST-elevation acute myocardial infarction. Heart Vessels. 2021;36:297–307.

29. Tsujita K, Sugiyama S, Sumida H, Shimomura H, Yamashita T, Yamanaga K, Komura N, Sakamoto K, Oka H, Nakao K, et al. Impact of Dual Lipid-Lowering Strategy With Ezetimibe and Atorvastatin on Coronary Plaque Regression in Patients With Percutaneous Coronary Intervention. J Am Coll Cardiol. 2015;66:495–507.

30. Wiviott SD, Braunwald E, McCabe CH, Montalescot G, Ruzyllo W, Gottlieb S, Neumann F-J, Ardissino D, De Servi S, Murphy SA, et al. Prasugrel versus Clopidogrel in Patients with Acute Coronary Syndromes. New England Journal of Medicine. 2007;357:2001–2015.

31. Levine GN, Jeong Y-H, Goto S, Anderson JL, Huo Y, Mega JL, Taubert K, Smith SC. World Heart Federation expert consensus statement on antiplatelet therapy in East Asian patients with ACS or undergoing PCI. Nat Rev Cardiol. 2014;11:597–606.

